# The comparison of vaccine hesitancy of COVID-19 vaccination in China and the United States

**DOI:** 10.1101/2021.04.29.21256317

**Authors:** Taoran Liu, Zonglin He, Jian Huang, Ni Yan, Qian Chen, Fengqiu Huang, Yuejia Zhang, Omolola M Akinwunmi, Babatunde Akinwunmi, Casper J.P Zhang, Yibo Wu, Wai-Kit Ming

## Abstract

**Objectives:** To investigate the differences in vaccine hesitancy and preference of the currently available COVID-19 vaccines between two countries, viz. China and the United States (US).

**Method:** A cross-national survey was conducted in both China and the US, and discrete choice experiments as well as Likert scales were utilized to assess vaccine preference and the underlying factors contributing to the vaccination acceptance. A propensity score matching (PSM) was performed to enable a direct comparison between the two countries.

**Results:** A total of 9,077 (5,375 and 3,702, respectively, from China and the US) respondents have completed the survey. After propensity score matching, over 82.0% respondents from China positively accept the COVID-19 vaccination, while 72.2% respondents form the US positively accept it. Specifically, only 31.9% of Chinese respondents were recommended by a doctor to have COVID-19 vaccination, while more than half of the US respondents were recommended by a doctor (50.2%), local health board (59.4%), or friends and families (64.8%). The discrete choice experiments revealed that respondents from the US attached the greatest importance to the efficacy of COVID-19 vaccines (44.41%), followed by the cost of vaccination (29.57%), whereas those from China held a different viewpoint that the cost of vaccination covers the largest proportion in their trade-off (30.66%), and efficacy ranked as the second most important attribute (26.34%). Also, respondents from China tend to concerned much more about the adverse effect of vaccination (19.68% vs 6.12%) and have lower perceived severity of being infected with COVID-19.

**Conclusion:** While the overall acceptance and hesitancy of COVID-19 vaccination in both countries are high, underpinned distinctions between countries are observed. Owing to the differences in COVID-19 incidence rates, cultural backgrounds, and the availability of specific COVID-19 vaccines in two countries, the vaccine rollout strategies should be nation-dependent.

## Introduction

A pneumonia-like disease outbreak, now named the 2019 Coronavirus disease (COVID-19) caused by a newly identified coronavirus, a.k.a., the severe acute respiratory syndrome (SARS)-CoV-2, has swept the globe in early 2020^1^. Although its exact origin remains unknown, and detailed knowledge of its transmission is still limited, this global pandemic has become the most serious public health threat from a respiratory virus since the 1918 H1N1 influenza pandemic^2-4^. One year after the commencement of the pandemic^5^, the COVID-19 is continuously imposing tremendous burdens on the public health systems and the economy globally^6,7^. As of early March 2021, 219 countries or regions have reported confirmed cases^8^, with over 120 million confirmed cases and over 2·66 million deaths, and a case fatality rate over 2·21% worldwide^9^.

Various public health measures, specifically, non-pharmaceutical interventions (NPIs) that showed effectiveness in previous infection outbreaks (e.g., mass facemask use, social distancing, and home quarantine) have been implemented by governments to contain the spread of SARS-CoV-2^2,10-12^. While the effectiveness of these public health measures in this outbreak remains to be determined, these NPIs may carry a high economic cost, leading to a long war of attrition in society^13^.

Therefore, massive vaccination coverage is considered as a prerequisite to achieve herd immunity and therefore curb the COVID-19 pandemic^14^. By March 2021, several COVID-19 vaccines^15-17^ have been developed and vaccination rollout has started in a few countries. However, people remain uncertain of the safety and efficacy of the vaccines. This becomes extensive especially after reports of adverse events secondary to shots of the mRNA vaccination^18^, sudden death secondary to shots of mRNA COVID-19 vaccines in Norway^19^, and incidences of inactivated vaccines during a phase 3 trial in Brazil, among others^20,21^. Also, according to a national poll in the US, only 58% of adults aged from 50 to 80 are willing to receive COVID-19 vaccine^22^. Additionally, as the increasing number of variants have been identified around the globe^23,24^, the current vaccines may no longer provide effective protection against SARS-CoV-2 virus including the variants.

Some studies have found that COVID-19 vaccines acceptance varied to a large extent across countries and regions before the vaccine becomes available, and the vaccine hesitancy of COVID-19 is increasing globally^25,26^. This may influence the vaccination coverage and hinder the establishment of herd immunity. Interestingly, an increasing vaccine hesitancy is being noticed among Chinese residents and some Asian countries where the transmission of COVID-19 has been well-controlled, specifically, when the vaccines are completely free in some countries, and such phenomena may be attributable to the low perceived benefits of the vaccines and low perceived risks of the COVID-19 pandemic, and most importantly, the low perceived efficacy and the increasingly high skepticism of the efficacy and safety of the currently COVID-19 vaccines^27^. Therefore, it is urgently to find the hindering factors and further promote the COVID-19 vaccination to achieve the herd immunity.

Vaccine hesitancy is multifactorial, attributable to general misinformation spread, perceived risks of disease, perceived efficacy and safety of vaccines, attitudes and demand of vaccines, cultural and religious factors and other unspecific factors^28,29^. China and the United States (US) are two of the representative countries that are among the major countries hardest hit by the pandemic^30^, and both have large population but very different cultural and religious systems^31^; hence residents’ acceptance to COVID-19 vaccination of the two countries could be strategically significant for further vaccines promotion and herd immunity.

Nevertheless, the direct comparison between countries could be biased and irrelevant owing to the high heterogeneity and confounding effects, hence in the present study, the propensity score matching (PSM) was utilized to compare the two population. And the following aims were addressed: 1) to investigate on the census-level acceptance and preference of residents and further exploring the influencing factors underlying their decision-making towards COVID-19 vaccination; 2) to compare the two countries in a statistically comparable way to demonstrate the vaccine hesitancy in two countries and further provide insights into future strategies of the large-scale COVID-19 vaccination coverage in two countries.

## Materials and Methods

### Study design

An anonymous self-administered cross-sectional survey was conducted online in China and the US through multiple international online panel providers (for data collecting in the US) and recruited volunteers across China (for data collecting in China), and stratified sampling was used, and nationally representative samples of the general adult populations were collected^32^. The questionnaire was established in Lighthouse studio (Sawtooth Software, version 9.8.1). The study was approved by the Jinan University Institutional Review Board. In the survey, respondents responded to a total of 55 items of questions, including standard demographic and socio-economic questions including age, sex, level of education, annual income, and marital status, followed by one set of discrete choice experiments (DCE) and questions about risk perception, COVID-19 impact, attitudes, and acceptance of and attitudes towards vaccines against COVID-10 during the pandemic. Prior to formal data collection, a pilot study in China had been conducted to evaluate the content validity and the reliability of the questions with both experts and general populations, and a group of experts was consulted to improve semantics and readability.

### Respondents

The inclusion criteria were respondents aged 18 years and above without cognitive impairments (self-report). Respondents were randomly recruited and selected through multiple international online panel providers (MTurk and Dynata) and by nearly 100 experienced volunteers recruited across China using a stratified sampling method. No personally identifiable information was collected. A total of 12,959 respondents was recruited, with a total of 9077 respondents (5,375 and 3,702 respondents from China and the US respectively) included in our study.

### Data collection

The survey data were collected between January 29 and February 13, 2021. The respondents were required to provide a randomly generated code after completing the survey to ensure that the respondents are real persons instead of robots. All the questions were close ended, with tick boxes provided for responses and no question skipping allowed, and no data would be stored if the website of the questionnaire was closed before completion. Hence, no missing data were generated.

General acceptance was defined as scoring greater than 6 with regards to answering the question “How do you rate your willingness and acceptance to get COVID-19 vaccination? (if the vaccines are generally available)”, and the *acceptance under social cues* was defined as scoring greater than 6 for the acceptance for COVID-19 vaccines if the vaccination was recommended by the respondents’ family members, friends or employers. Countries were categorized as developed and developing countries according to United Nations country classification^2^. Educational level was further classified into four groups, where “low” signifies respondents reporting having not finished a secondary education (high school); “medium” signifies those who had completed secondary, vocational, or equivalent degrees; and “high” group consisted of those who had completed a tertiary or bachelor’s degree and “very high” indicates postgraduate work. Moreover, two external COVID-19-related variables were added abided by the Method reported by Lazarus et al., namely the total SARS-CoV-2 positive cases per million persons and total SARS-CoV-2 deaths per million persons as reported by Worldometer on 21 January 2021^1, 2^. For COVID-19 cases per million in population, “High” was defined as more than 10,000 cases per million people, “medium” as between 1,000 and 10,000 cases per million people, and “low” was defined as below 1,000 cases per million people. For COVID-19-specific mortality cases per million in the population, “High” was defined as more than 1,000 deaths per million people, “medium” as between 100 and 1,000 deaths per million people, and “low” was defined as below 100 deaths per million people.

### Survey design

The questionnaire consists of three sections with a total of 55 items that required a response. In the first section, respondents responded to provide social-demographic information regarding age, sex, educational level, occupation, income level, nationality and marital status. Additionally, respondents were required to rate their willingness and acceptance to get vaccinated from “totally unwilling” to “totally willing” with and without social cues. And the questions were “How do you rate your willingness and acceptance of getting vaccination?” and “How do you rate your willingness and acceptance if your friends, family members, neighbors etc. recommend you do so?”, respectively. Also, respondents responded to questions related to COVID-19 infection history as well as major source of information with regard to COVID-19 vaccines.

In the second section, DCE was used to further explore respondents’ preference of the currently available vaccines. Specifically, vaccine attributes and their levels were identified and retrieved through relevant literature and several vaccines on the market, and the attributes were then ranked, categorized and refined by a group of experts in the field of public health and vaccination^3, 4^. A total of six attributes were identified and a two-vaccine profile was randomly adopted for the DCE design. The detailed description of the attributes and levels are summarized in Table S2. During the survey, respondents were asked to make a series of hypothetical choices and estimate their preference for different attributes of the vaccine based on scenarios.

In addition, to assess respondents’ attitudes toward COVID-19 and the acceptance of COVID-19 vaccination and their influencing factors, questions were designed based on the five-concept health belief model and other frameworks, as reported by various previous studies to assess vaccine acceptance and hesitancy for newly emerging infectious diseases such as H1N1, MERS or Ebola ^5^ (Figure S5). The following contents were included in this section: (1) the perceived susceptibility to COVID-19 infection; (2) perceived severity of COVID-19 infection, (3) perceived benefits of COVID-19 vaccination; (4) perceived barriers to COVID-19 vaccination and (5) cues to action for COVID-19 vaccination, such as a recommendation from a doctor or local health board, were assessed; (6) socio-economic factors; and (7) past immunization behaviors ^6^. Most questions were assessed on a seven-point Likert scale. The reliability (alpha = 0·8951) and validity (Kaiser-Meyer-Olkin Measure = 0·942) of the Likert scale were tested.

### Statistical analysis

The data in this study were collected in an international cross-sectional survey. To minimize potential confounding biases due to discrepancy in baseline characteristics, propensity scoring was calculated and matched to balance covariates for respondents in China and in the United States. Propensity score matching is a statistical technique that can help strengthen causal arguments in quasi-experimental and observational studies by reducing selection bias^33^. The covariates were identified using the pair-wise Pearson correlation matrix, and the results were presented in the Table S3. Later, the set of covariates was determined by at best minimizing the residual confounding factors, where a logistic regression model was performed to estimate the propensity scores for each groups of respondents, and the covariate imbalance testing was reported in the Table S5. Finally, a total of 3,436 respondents, with half from China and the other half from the US, were matched using propensity scores from the total 12,959 respondents, with the covariates being sex, age, and annual income.

Descriptive statistics were performed to describe the characteristics of socio-economic status and demographic information, risk perception, pandemic impact, as well as acceptance, attitudes and preferences of COVID-19 vaccines, using central tendency (mean, median) and dispersion (standard deviation, interquartile interval) measures. The Chi-square test or Fischer’s exact test was used for the univariate analysis of qualitative variables and Student’s t-test or Mann Whitney test for quantitative variables. Absolute and relative frequencies were presented for qualitative variables, while quantitative variables were presented as mean (standard deviation). Multivariate logistic regression was then performed between the vaccine demand group and vaccine delay group to identify the influencing factors of vaccination acceptance (immediate or delayed acceptance), with the odds ratio (OR), standard error (SE), and a 95% confidence interval (CI) being calculated. The data were analyzed using STATA, version 14·0 (Stata Corp, College Station, TX, USA). For the DCE part, we performed a conditional logit model (CLOGIT) to quantify respondents’ preference for vaccines’ attributes and levels in their trade-off in general and to further explore participants’ preference heterogeneity among different countries and regions. After the conditional logit model, we dummy coded all the attribute levels with the level least preferred selected as the reference level^7^. After dummy coding, the model parameter β represents the value that respondents placed on an attribute level relative to the reference level, where the model parameter β does not directly reflect the preference weight within an attribute this presentation can enhance the interpretation of the preference weights specifying the difference between two random coefficients. The data of the DCE part were analyzed using STATA, version 14·0 (Stata Corp, College Station, TX, USA).

## Scenario analysis and simulation

We also performed a scenario analysis and product simulation in Lighthouse studio (version 9·9·1) to further explore vaccines with what characteristics most contributed to respondents’ preference and with most probability to be uptake. The base scenario was hypothesized based on the vaccines variety with every attributes’ levels being the lowest utilities (except cost attribute), while the best scenario was assumed to have each attributes’ levels being with the highest utilities (except cost attributes). And the other scenarios were established according to the currently available information of vaccines from various clinical trials. Also, we use share of preference as our simulation model since this model helped us better predict the level of preference any vaccines might achieve. And the whole simulation were done in two steps: 1) Subject the respondent’s total utilities for the product to the exponential transformation, specifically as *s* = *exp*(*utility*), and 2) rescale the results to a total of 100%^34^.

## Results

### Respondents’ characteristics

The present study conducted a large-scale self-administered online survey in China and US. A total of 9077 respondents (5,375 and 3,702 respondents from China and the US respectively), who have completed the survey, were selected and further analyzed in the present study. Concerning the pre-PSM samples, the respondents from the United States tended to be higher-educated and earning more money annually compared to respondents from China. For the two respondent groups, the majority of the respondents were female (55·3% for China, 51·8% for the US), and 47·8% of the respondents in China and 67·6% of those in the US hold a bachelors’ degree or higher. After PSM for age, sex, education, annual income, and occupation, no statistically significant discrepancies could be discerned between the respondents from the two countries in demographic characteristics (*P* = 1·00 for age, sex, education, annual income and occupation).

### Generate hesitancy and participants’ vaccination history

As pre-PSM listed in Table 1 and Fig 2, respondents from China showed a relatively lower hesitancy of COVID-19 vaccines (7·8/10) than those from the US (7·2/10) when asked “How do you rate your willingness and acceptance to get COVID-19 vaccination? (if the vaccines are generally available)” (*general acceptance*). For post-PSM, over 82·0% respondents from China positively accept the COVID-19 vaccination, while 72·2% respondents form the US positively accept it, and the proportion change to 83·3% and 71·0%, respectively, if the respondents were recommended to get vaccinated by friends, family members or employers, etc. Additionally, around 12% respondents from China and 38·3% from the US have ever delayed or cancelled vaccination for reasons other than illness or allergy, respectively.

**Table 1.**
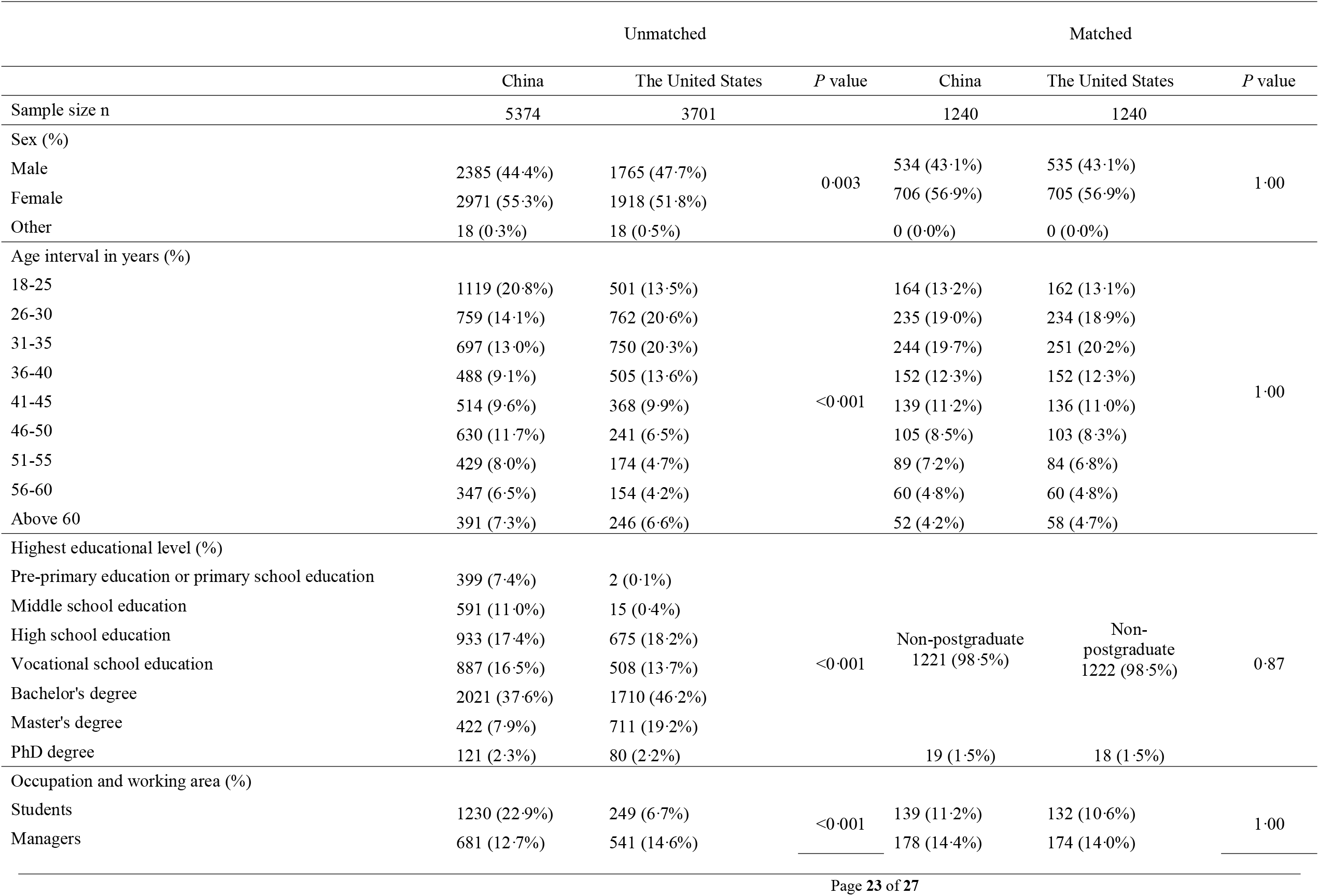
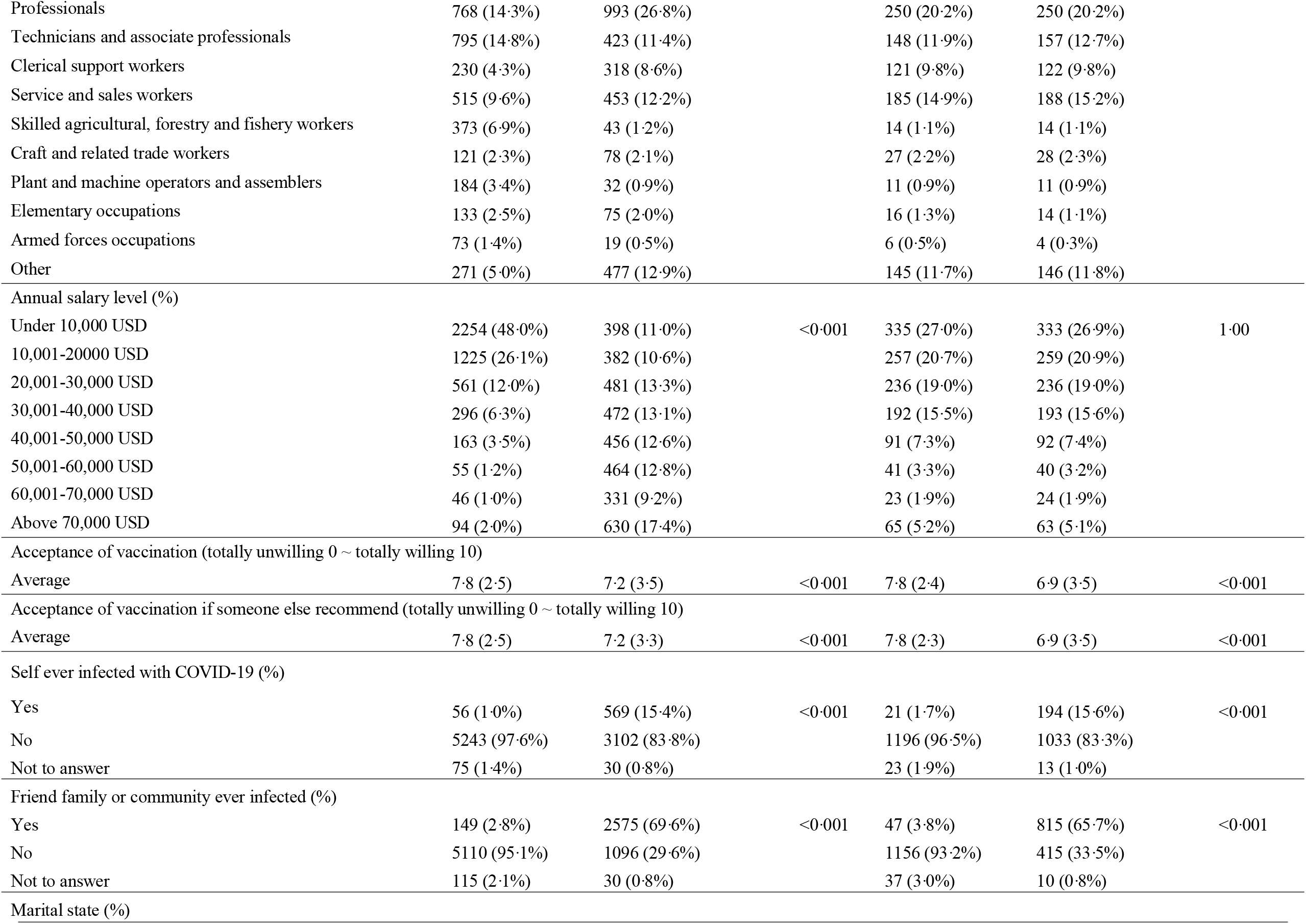
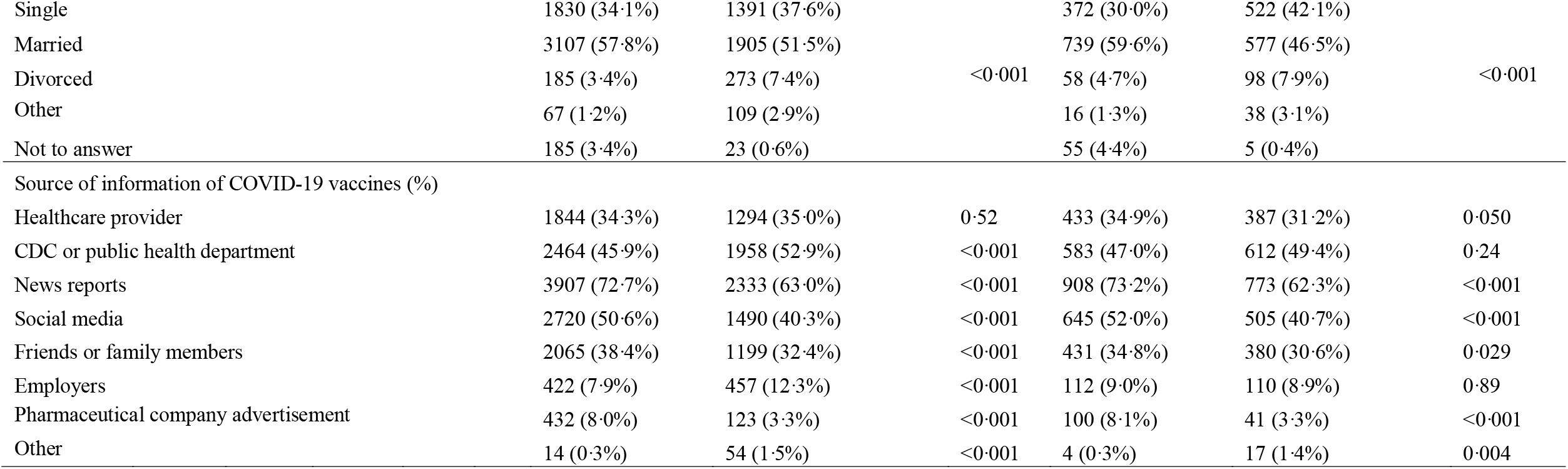
Participants’ demographic information, major source of information and acceptance.

**Fig 1.**
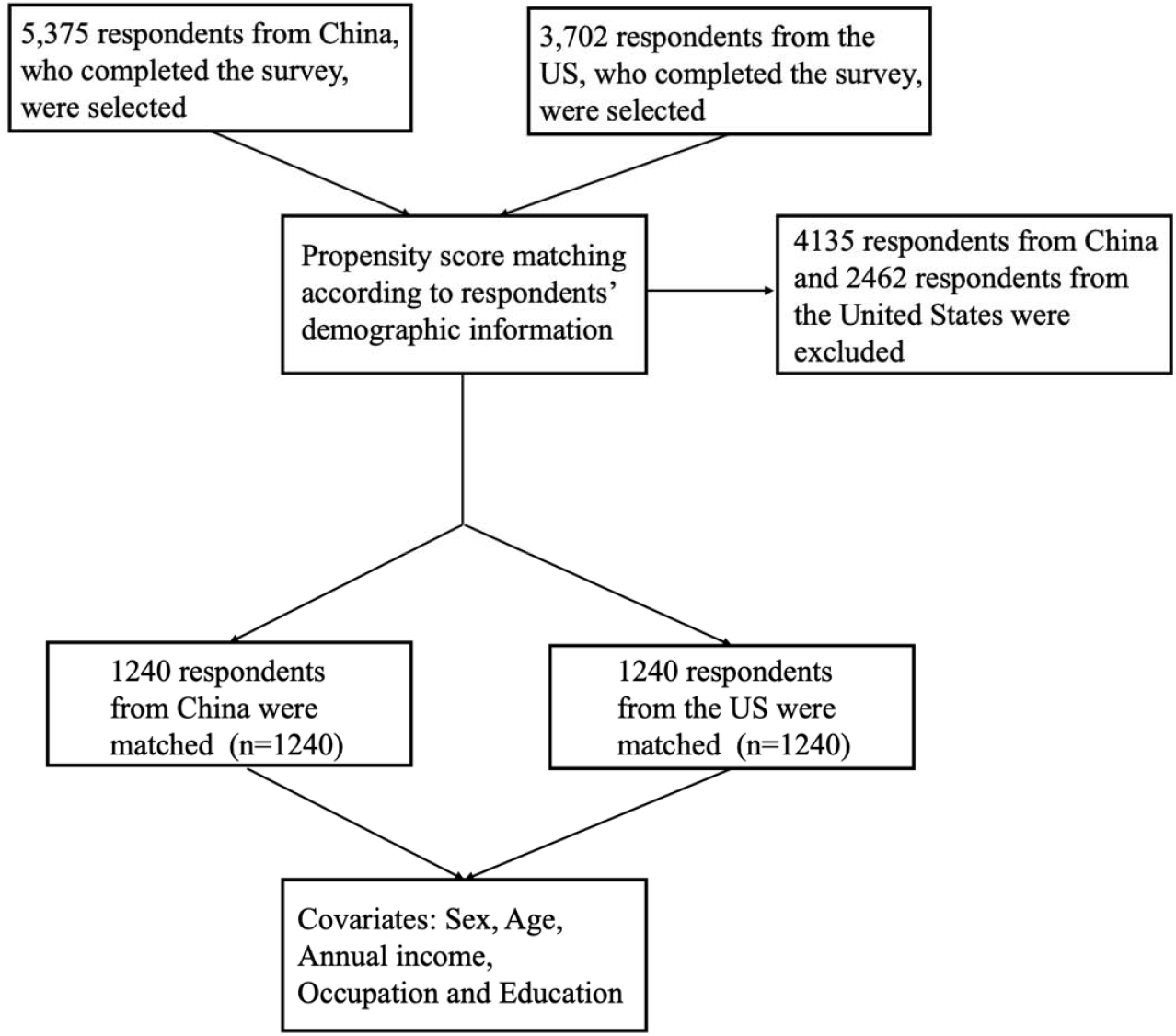
The flow chart of propensity score matching.

**Fig 2.**
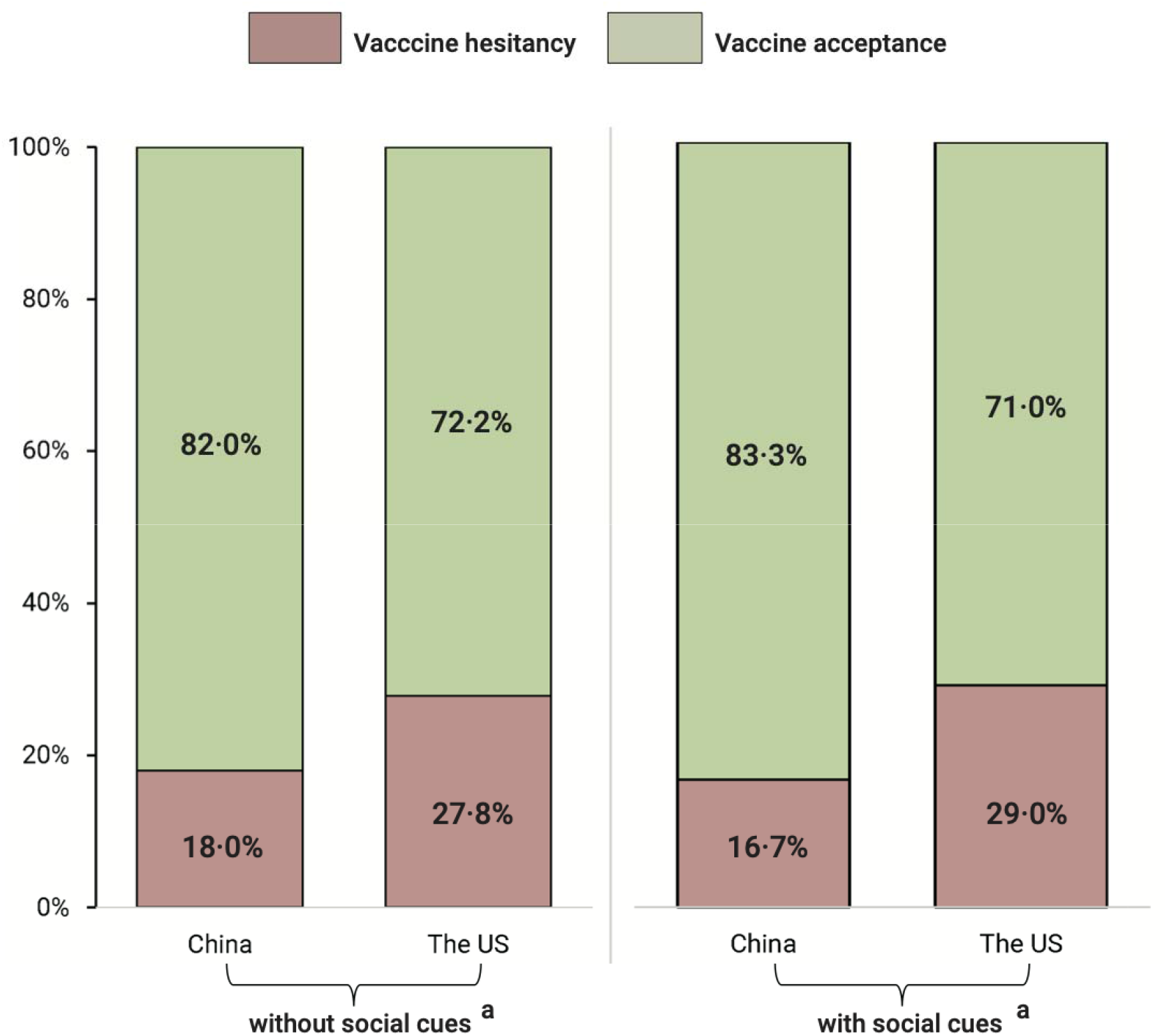
COVID-19 vaccination acceptance comparison between China and the US after propensity score matching. ^a^ Social cues in this study means social factors that potentially impact respondents’ acceptance, i.e., recommendations from friends, family members, or employers, etc.

Interestingly, for post-PSM, 31·9% of Chinese respondents were recommended by a doctor to get COVID-19 vaccination, while more than half of the US respondents (50·2%) were recommended by a doctor (Table S1). And generally, more than half of the US respondents were recommended to get vaccination, either by local health board (59·4%), or by friends or families (64·8%).

The influence of demographic factors on the average rating of hesitancy of COVID-19 vaccination between the two countries for were shown in Table S1, and the detailed subgroup analysis of the hesitancy by sex, age intervals, education, occupation and annual income were shown in Figure S1-S5, respectively. Also, proportion of vaccine hesitancy from China generally smaller than those from the US, except for the respondents with a master’s academic degree, or with an annual income over 70,000 USD, or with a skilled, agricultural, forestry and fishery related occupations. In both China and the US, respondents’ vaccination hesitancy increased with the academic degree from bachelor’s degree. Moreover, being female and higher educated, as well as themselves being infected or friends or families around being infected served as contributor for the respondents hesitating the vaccines, as shown in Table S1.

### Post-PSM participants’ vaccine preference, attributes and level importance

The relative attributes’ importance compared between the United States and China was shown in Figure 3. After PSM, we found respondents from the US attached the greatest importance to the efficacy of COVID-19 vaccines (44·41%), followed by the cost of vaccination (29·57%), whereas those from China hold a different viewpoint, the cost of vaccination covers the largest proportion in their trade-off (30·66%), and efficacy ranked as the second most important attribute (26·34%). Additionally, respondents from China also concerned much more about the adverse effects of the vaccines, which was ranked as the third most important factor (19·68%). The duration of vaccination, vaccine varieties and time for the vaccine starting to work remained a relative low importance in both countries.

**Fig 3.**
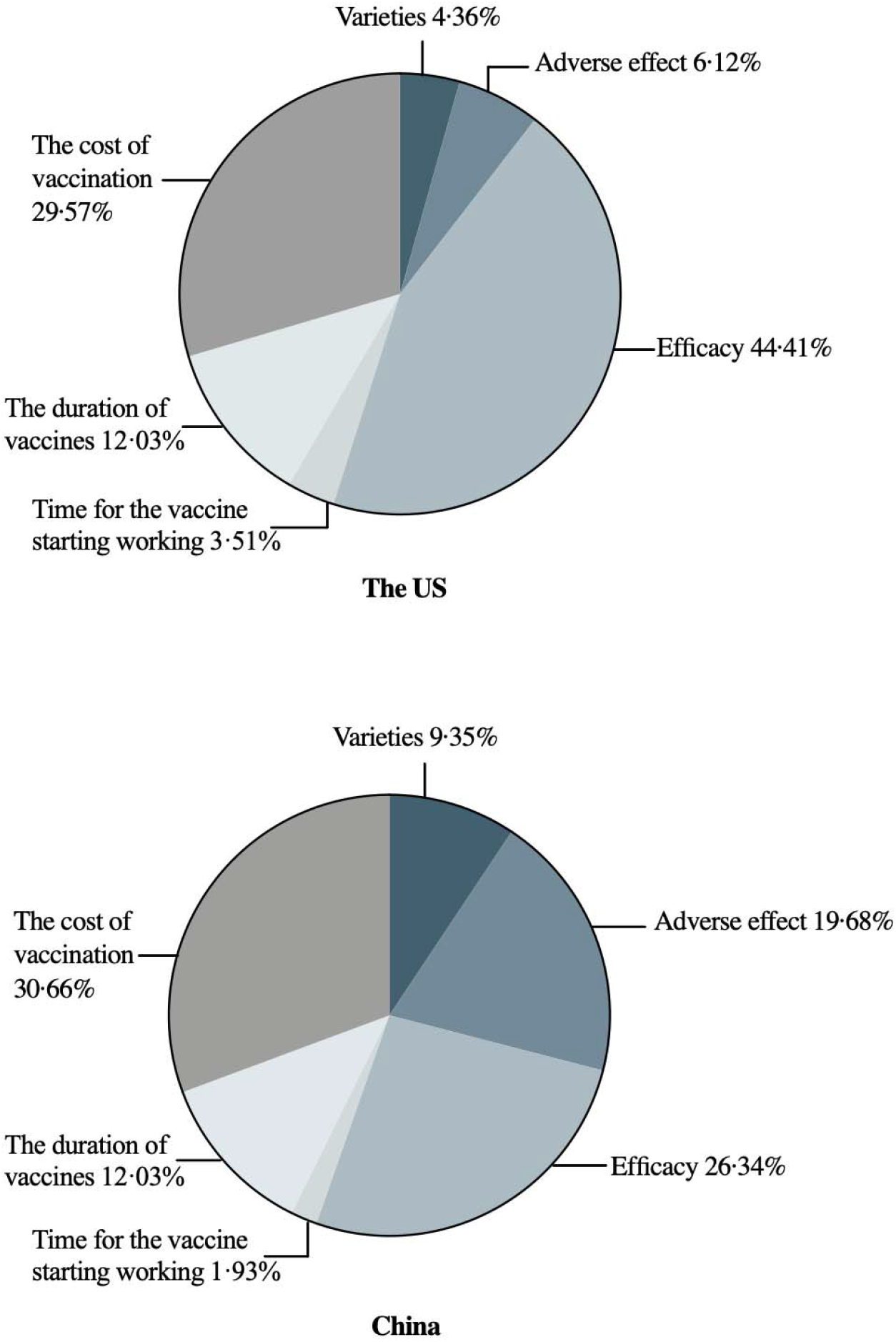
The relative importance of COVID-19 vaccines attributes comparison between respondents from the US and China.

Interestingly, in post-PSM result, respondents from the US preferred the mRNA COVID-19 vaccine, while respondents from China prefer the inactivated COVID-19 vaccine (vs mRNA) (OR=1·164, 95% CI (1·124, 1·205), p < 0·001). Also, respondents’ vaccine preference significantly decreased with a moderate adverse effect compared with a very mild adverse effect. In both countries, respondents’ preference increased along with the rise of efficacy, and reached a peak at 95% efficacy (vs 55%). The reduction of vaccine preference only appeared in respondents from the US if the time for the vaccines starting to work getting longer (20 days vs 5 days) (OR=0·893, 95% CI (0·849, 0·938), p =0·06). Moreover, respondents from both countries preferred a longer vaccine protection time and low level of vaccination cost.

The post-PSM preference comparison between sex in both countries is showed in Figure S6. For respondents from the US, first, both male and female respondents ranked the efficacy of COVID-19 vaccines as the most essential attribute, and the efficacy was even slightly more important for female (male: 43·12%, female 45·07%). while for respondents from China, the cost of vaccination had the greatest relative importance, and slightly more important for female (male: 28·85%, female: 31·45%). Similarly, the efficacy of COVID-19 vaccines also accounted more for female respondents than male respondents (male: 26·36%, female: 26·02%).

### Post-PSM scenario analysis and uptake likelihood prediction

Table 3 presented the simulated share of preference under nine different scenarios which were based on clinical trials and real-world data of various COVID-19 vaccines, as reported by various large-scale clinical trials. The base scenario is the vaccine with seemingly lower preference, and only 2·9% of the respondents from the US will uptake the base scenario vaccine, while 4·5% of those from China prefers to taking the base scenario vaccine. Still, we noticed that the US respondents are more likely to choose mRNA vaccines (share of preference 14·2% for Scenario 2 and 17·3% for Scenario 3) while those from China places higher preference in Inactivated vaccine (Scenario 4, share of preference 13·7%). But the respondents from the US also have high share of preference for Adenovirus vaccine (Scenario 5) and the Chinese respondents have high preference for mRNA vaccine too (Scenario 3, Share of preference 13·4%). If all the attributes of the vaccine were set as the best levels as indicated in the MNL analysis (Scenario 8), then 19·1% and 17·6% of the respondents from the US and China respectively will choose this hypothetical vaccine.

### Behavioral and psychological results

The Likert Scale, as presented in Table S1, indicated that generally, the respondents had a highly positive attitude towards the benefits of COVID-19 vaccines (the US: 15·8/21; China: 17/21) and were not so concerned about the risks and barriers of COVID-19 vaccination, although respondents from China concerned slightly more about risks than those from the US (the US: 11·3/21; China: 13·3/21). Also, the respondents generally believe in the necessity and efficacy of vaccination in the prevention of diseases, as they rate high on the item “In general, vaccination is effective in preventing diseases” (the US: 5·5/7·0; China: 5·7/7·0), followed by the scoring of 5·9/7.0 and 5·3/7·0 for respondents from China and the US respectively, for item “In general, prevention is better than cure”. In terms of socio-cultural factors, religion- and gender-related reasons were the least contributory to the decision-making of COVID-19 vaccination (the US: 3·1/7·0; China:4·2/7·0) for religious or cultural reasons, and (the US 4·4/7·0; China: 5·7/7·0 for gender reasons). Respondents rate the lowest score on the item “I believe that people are risking their health or the health of the society if they do not take a COVID-19 vaccine” (the US: 2·5/7·0; China: 3·5/7·0).

## Discussion

The present study sought to provide a comprehensive investigation of the acceptance of COVID-19 vaccination in China and the United States and compare the hesitancy and preference of COVID-19 vaccines between the two countries controlling for demographic characteristics using PSM. The unmatched samples of the respondents from the two countries were adjusted with uncontrolled quota sampling method, hence the results may demonstrate the census-level investigation in two countries. Generally, majority of the respondents in both countries had high acceptance regarding the COVID-19 vaccination, either general acceptance or acceptance under recommendation from friends, family or employers.

In our study, we found that respondents from China have a relatively higher acceptance than those from the US, and when compared to the studies conducted before the vaccine become available or even before the results of clinical trials on some COVID-19 vaccines were available, their acceptance increased respectively^35^. Existing opinion polls for acceptance of a hypothetical COVID-19 vaccines of US citizen ranged from 40% to 70%^36-39^. And a population-based survey conducted in Hong Kong, China reported the overall vaccine acceptance rate was only 37·2%^40^, while more than 90% of respondents in mainland China would like to be vaccinated for COVID-19 vaccines regardless of the efficacy, among whom^35^. Although studies have reported that some perceived susceptibility to infection or perceived risks may not be associated with the COVID-19 acceptance^40^, our results showed that there is distinct difference between the respondents from China and the US regarding the perceived risks of infection and perceived benefits of vaccination after propensity score matching. The Chinese respondents tended to be more concerned about the health problems COVID-19 may cause to them if they were infected, while the US respondents are more concerned of being infected (Table S1). And studies have found an association between the incidence of diseased population and perceived risks, and low risk perception was also associated with the respondents trusting health professionals and health officials for information on COVID-19^41^. Public concern about vaccine safety and efficacy has frequently been reported as one of the major obstacles to vaccination acceptance, specifically when the COVID-19 vaccines were being developed and rolled at an unprecedented speed^42-47^. But it is noteworthy that, despite high acceptance of the vaccines, Chinese respondents were more concerned about the safety of the vaccines, the adverse effect of the COVID-19 vaccines had a far more relative importance for respondents from China than those from the US. And combined with their low perceived risks of susceptibility, this may hence lead to their placing greater importance for the attribute “cost” in the discrete choice experiment (Table 2 and Figure 2) and also may explain why the vaccination rate still remains a relatively low level in China, compared with some other countries in the world, such as the US, the UK, etc. As of March 31^st^, 2021, there are a total of 114·69 million doses of COVID-19 vaccination reported in China; however, the vaccination rate remains below 10%^48^. There is still a huge gap from achieving herd immunity, which requires 70% to 90% vaccination rate as some experts estimated^49,50^. In contrast, the vaccination rate in the US has reached 30% (at least one dose)^51^ with 103·35 million people receiving one or more doses cumulative. And therefore, although, vaccination growth rate seems to be more rapid than the US from March 24, 2021, the speed is not rapid enough to reach herd immunity in short term, eventually, the relative low level of vaccination rate in China may force China to lose its advantage in COVID-19 epidemic control^52^. Therefore, improving the speed of vaccination is a top priority currently for all countries around the world to terminate the pandemic.

In both pre-PSM and post-PSM samples, we found that while the respondents from China attached more importance to cost, those from US attached more to efficacy. This finding may strongly correlate with local COVID-19 incidence and mortality rate, namely the perceived risk for the public to be infected. As of March 14, 2021, there are over 120 million cumulative COVID-19 cases and around 2·66 cumulative death^9^ in the US. Thus, vaccines with high efficacy are urgently needed to contain the development of epidemic in the US, whereas in China, there are only more than 102 thousand cumulative cases and around 5 thousand deaths^9^. Also, we found that respondents from the US prefer to be vaccinated with mRNA vaccines, while respondents from China consider inactivated vaccines as the best choice, this might be related to the actual availability of different types of vaccine in US and China. High vaccination coverage depends on the public understanding of the need and value of vaccination, the availability of vaccines as well as accessible immunization services ^53^. And to further promote high vaccination coverage, the Strategic Advisory Group of Experts (SAGE) on Immunization Working Group on Vaccine Hesitancy recommends three categories of strategies, namely the increase of understanding of vaccine hesitancy, establishment of structures and organizational capacity at global, national and local levels, as well as international collaboration between countries regarding the development, validation, and implementation of new tools to address hesitancy ^53,54^. Also, our study emphasizes the key role of international collaboration. Humans have been fighting for the many centuries, and the formal collaboration has been institutionalized through World Health Organization (WHO). However, in this COVID-19 pandemic, many countries showed poor performance against COVID-19 with refection of strong self-interested nationalism^55^. The nationalism has greatly restricted international collaboration, and further hindered the epidemic control. Meanwhile, vaccine nationalism has been warned by WHO as a moral failure. Therefore, to relieve the global epidemic, collaboration on vaccine were urgently needed, especially when some adenovirus COVID-19 vaccination have been suspended in many countries due to severe adverse effects^56^.

**Table 2.** Comparison between the US and China in attributes levels utility and odds ratios·.

| Attributes | Variable | The US (n=1240) |  |  |  |  | China (n=1240) |  |  |  |  |
| --- | --- | --- | --- | --- | --- | --- | --- | --- | --- | --- | --- |
|  |  | Coefficient | Std Error | OR | 95% CI | P value | Coefficient | Std Error | OR | 95% CI | P value |
| Varieties | mRNA | Reference |  |  |  |  |  |  |  |  |  |
|  | Adenovirus vector vaccines | 0.006 | 0.019 | 0.940 | (0.906 - 0.976) | 0.761 | -0.105 | 0.018 | 0.922 | (0.890 - 0.954) | <0.001 |
|  | Inactivated vaccine | -0.073 | 0.019 | 0.869 | (0.837 - 0.902) | <0.001 | 0.128 | 0.018 | 1.164 | (1.124 - 1.205) | <0.001 |
| Adverse effect | very mild | Reference |  |  |  |  |  |  |  |  |  |
|  | mild | 0.070 | 0.019 | 1.013 | (0.976 - 1.051) | <0.001 | 0.077 | 0.018 | 0.879 | (0.849 - 0.910) | <0.001 |
|  | moderate | -0.128 | 0.019 | 0.831 | (0.800 - 0.863) | <0.001 | -0.284 | 0.018 | 0.612 | (0.591 - 0.635) | <0.001 |
| Efficacy | 55% | Reference |  |  |  |  |  |  |  |  |  |
|  | 65% | -0.516 | 0.028 | 1.213 | (1.147 - 1.282) | <0.001 | -0.246 | 0.025 | 1.088 | (1.036 - 1.144) | <0.001 |
|  | 75% | 0.061 | 0.029 | 2.158 | (2.038 - 2.285) | 0.038 | 0.048 | 0.028 | 1.461 | (1.384 - 1.543) | 0.084 |
|  | 85% | 0.438 | 0.025 | 3.146 | (2.994 - 3.306) | <0.001 | 0.204 | 0.024 | 1.707 | (1.627 - 1.791) | <0.001 |
|  | 95% | 0.726 | 0.027 | 4.196 | (3.980 - 4.424) | <0.001 | 0.326 | 0.026 | 1.928 | (1.832 - 2.030) | <0.001 |
| Time for the vaccine starting working | 5 days | Reference |  |  |  |  |  |  |  |  |  |
|  | 10 days | -0.035 | 0.024 | 0.904 | (0.863 - 0.948) | 0.142 | 0.021 | 0.022 | 1.025 | (0.982 - 1.069) | 0.324 |
|  | 15 days | 0.017 | 0.023 | 0.953 | (0.912 - 0.996) | 0.445 | -0.027 | 0.022 | 0.976 | (0.936 - 1.019) | 0.217 |
|  | 20 days | -0.048 | 0.025 | 0.893 | (0.849 - 0.938) | 0.060 | 0.008 | 0.024 | 1.011 | (0.965 - 1.059) | 0.732 |
| The duration of vaccine works | 5 months | Reference |  |  |  |  |  |  |  |  |  |
|  | 10 months | -0.081 | 0.024 | 1.148 | (1.096 - 1.202) | <0.001 | -0.021 | 0.022 | 1.153 | (1.105 - 1.204) | 0.338 |
|  | 15 months | 0.131 | 0.024 | 1.419 | (1.354 - 1.487) | <0.001 | 0.048 | 0.023 | 1.236 | (1.182 - 1.292) | 0.035 |
|  | 20 months | 0.169 | 0.023 | 1.475 | (1.410 - 1.543) | <0.001 | 0.136 | 0.022 | 1.350 | (1.293 - 1.409) | <0.001 |
| The cost of vaccination | \$0 | Reference | | | | | | | | | |
| | \$50 | 0.179 | 0.026 | 0.674 | (0.640 - 0.709) | <0.001 | 0.162 | 0.025 | 0.776 | (0.740 - 0.814) | <0.001 |
| | \$100 | -0.111 | 0.026 | 0.504 | (0.479 - 0.530) | <0.001 | -0.029 | 0.024 | 0.641 | (0.612 - 0.672) | 0.225 |
| | \$150 | -0.263 | 0.029 | 0.433 | (0.409 - 0.458) | <0.001 | -0.200 | 0.027 | 0.540 | (0.513 - 0.570) | <0.001 |
| | \$200 | -0.381 | 0.031 | 0.385 | (0.362 - 0.409) | <0.001 | -0.349 | 0.029 | 0.466 | (0.440 - 0.493) | <0.001 |

**Table 3.** Share of preference and scenario analysis results.

| The US |  |  |  |  |  |  |  |  |  |
| --- | --- | --- | --- | --- | --- | --- | --- | --- | --- |
|  | Base scenario | Scenario 1 | Scenario 2 | Scenario 3 | Scenario 4 | Scenario 5 | Scenario 6 | Scenario 7 | Scenario 8 |
| Vaccine varieties | Inactivated vaccine | Adenovirus vaccine | mRNA | mRNA | Inactivated vaccine | Adenovirus vaccine | Adenovirus vaccine | Adenovirus vaccine | mRNA |
| Adverse effect | moderate | Very mild | Moderate | Mild | Very mild | Mild | Very mild | Very mild | Very mild |
| Efficacy | 55% | 75% | 95% | 95% | 75% | 95% | 65% | 75% | 95% |
| Time for the vaccine starting working | 20 days | 20 days | 20 days | 20 days | 10 days | 20 days | 5 days | 5 days | 5 days |
| The duration of vaccine works | 5 months | 5 months | 5 months | 5 months | 5 months | 5 months | 5 months | 5 months | 20 months |
| The cost of vaccination | \$50 | \$50 | \$50 | \$50 | \$50 | \$50 | \$50 | \$50 | \$50 |
| Share of preference | 2.9% | 8.2% | 14.2% | 17.3% | 7.7% | 16.2% | 5.2% | 9.2% | 19.1% |
| China |  |  |  |  |  |  |  |  |  |
| Vaccine varieties | Adenovirus vaccine | Adenovirus vaccine | mRNA vaccine | mRNA vaccine | Inactivated vaccine | Adenovirus vaccine | Adenovirus vaccine | Adenovirus vaccine | Inactivated vaccine |
| Adverse effect | moderate | Very mild | Moderate | Mild | Very mild | Mild | Very mild | Very mild | Very mild |
| Efficacy | 55% | 75% | 95% | 95% | 75% | 95% | 65% | 75% | 95% |
| Time for the vaccine starting working | 20 days | 20 days | 20 days | 20 days | 10 days | 20 days | 5 days | 5 days | 5 days |
| The duration of vaccine works | 5 months | 5 months | 5 months | 5 months | 5 months | 5 months | 5 months | 5 months | 20 months |
| The cost of vaccination | \$50 | \$50 | \$50 | \$50 | \$50 | \$50 | \$50 | \$50 | \$50 |
| Share of preference | 4.5% | 10.7% | 9.4% | 13.4% | 13.7% | 12.4% | 7.9% | 10.6% | 17.6% |

Our findings are useful for designing and modifying effective vaccination promotion strategies and immunization coverage programs for the public and those with vaccine hesitancy, based on different national conditions and contextual backgrounds. Firstly, the perceived risks for infections and risks for severe health problems secondary to the infection may be vital for the respondents actively to be vaccinated. Hence the risks for the infection and the adverse outcomes secondary to the infection should be clearly delivered by media or governments to the public to enhance mutual understanding. Secondly, the vaccine price should be affordable and available for the public, and it is auspicious that many countries have been making COVID-19 vaccines free of charge for the public^57-59^. Moreover, monitoring information about vaccine safety should be made public on a regular basis after the application of the vaccine, and timely health education and communication conducted by authoritative sources^44,60^.

Additionally, China and the US have plenty of differences, especially in contextual influences, for instance, media environment, influential leaders, historical influences, policies, cultural factors, etc. As the majority (82·0% for Chinese respondents, 72·2% for US respondents) of respondents generally accept the COVID-19 vaccination, it is still worth efforts to further identify other barriers or facilitators to their vaccination decision based on different contextual backgrounds and different countries.

Although the average COVID-19 vaccines acceptance of respondents from the US may be relatively lower than those from China, higher annual income was associated with higher vaccine hesitancy among US respondents. The respondents with higher educational level may tend to acknowledge more vaccines risks.

### Strength and limitations

The present study has limitations. The inherent nature of cross-sectional studies renders it difficult to establish causality or to generalize the results in a long-term manner, especially when vaccination acceptance is variable, dynamic and multifactorial. Hence the results of the study should be interpreted with cautions. But the present study implemented the PSM to minimize the confounding effects, and it may somehow enhance the interpretability of the results.

Still, the present study is the first study to directly compare the acceptance and preference of the respondents from two distinctly different representative countries, and the results of the study may provide insights for the vaccination promotion strategy based on different national situations globally. And the present study used multiple study methods to provide the most comprehensive and updated investigation, especially on the influencing factors contributing to the decision making about vaccination.

In conclusion, great variability in the preference of COVID-19 vaccines was found between respondents from China and the United States, and the influencing factors for the hesitancy as well as the attributes for their preferences for the vaccines varied, hence multi-disciplinary and international collaboration should be established and strengthened based on specific national conditions to further reduce vaccine hesitancy and increase vaccination coverage.

## Data Availability

Data can be achieved via emailing to

## Author contributions

Conceptualization: TRL, ZLH, WKM

Methodology: TRL, ZLH, JH, NY

Investigation: TRL, ZLH, QC, FQH, YZ, OMA, WKM, YBW

Visualization: TRL, ZLH, NY

Supervision: WKM, YBW

Writing—original draft: TRL, ZLH

Writing—review & editing: TRL, ZLH, JH, NY, BA, CJPZ

## Data and materials availability

Data can be achieved via emailing to

## Ethical approval

This study has received the ethical approval at Jinan University Medical Ethics Committee. The approval code is JNUKY-2021-004

## Supplementary Materials

**Fig S1. COVID-19 vaccination acceptance comparison between China and the US depending on sex. and age intervals after propensity score matching**

**Fig S2. COVID-19 vaccination acceptance comparison between China and the US depending on education and annual income after propensity score matching**

**Fig S3. COVID-19 vaccination acceptance comparison between China and the US depending on occupation after propensity score matching**

**Fig S4. COVID-19 vaccination acceptance in China before propensity score matching**

**Fig S5. COVID-19 vaccination acceptance in the US before propensity score matching**

**Fig S6. COVID-19 vaccination preference comparison between China and the US depending on sex after propensity score matching**

**Table S1. Behavioral and psychological results**

**Table S2. The attributes and levels of vaccine acceptance and preference**.

**Table S3. The Pearson Correlation matrix between the baseline characteristics and acceptance**

