## Supplementary material for "The comparison of vaccine hesitancy of COVID-19 vaccination in China and the United States": SUPP FINAL VERSION.pdf

### Supplementary information

**Fig S1. COVID-19 vaccination acceptance comparison between China and the US depending on sex and age intervals after propensity score matching**

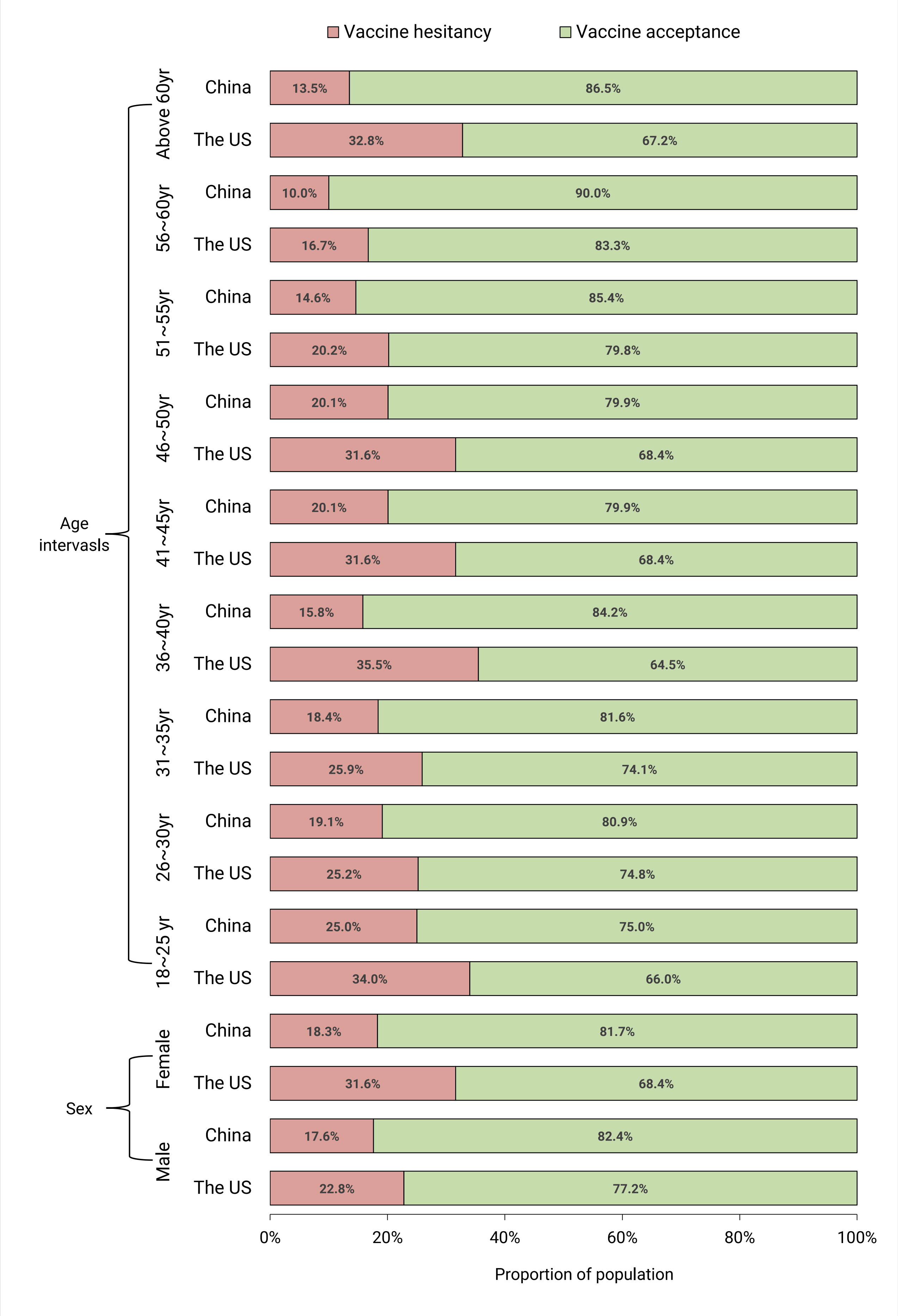

**Fig S2. COVID-19 vaccination acceptance comparison between China and the US depending on education and annual income after propensity score matching**

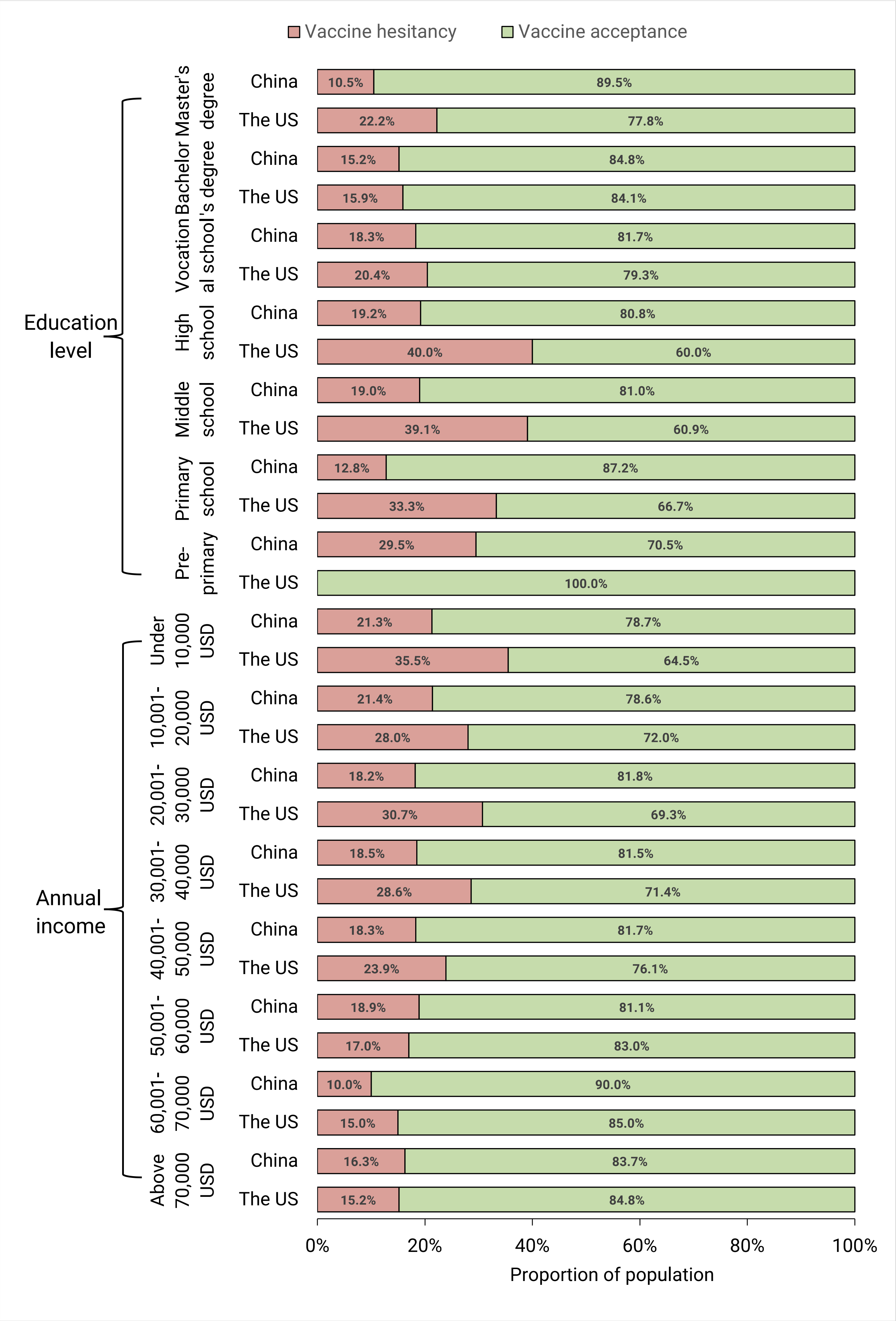

**Fig S3. COVID-19 vaccination acceptance comparison between China and the US depending on occupation after propensity score matching**

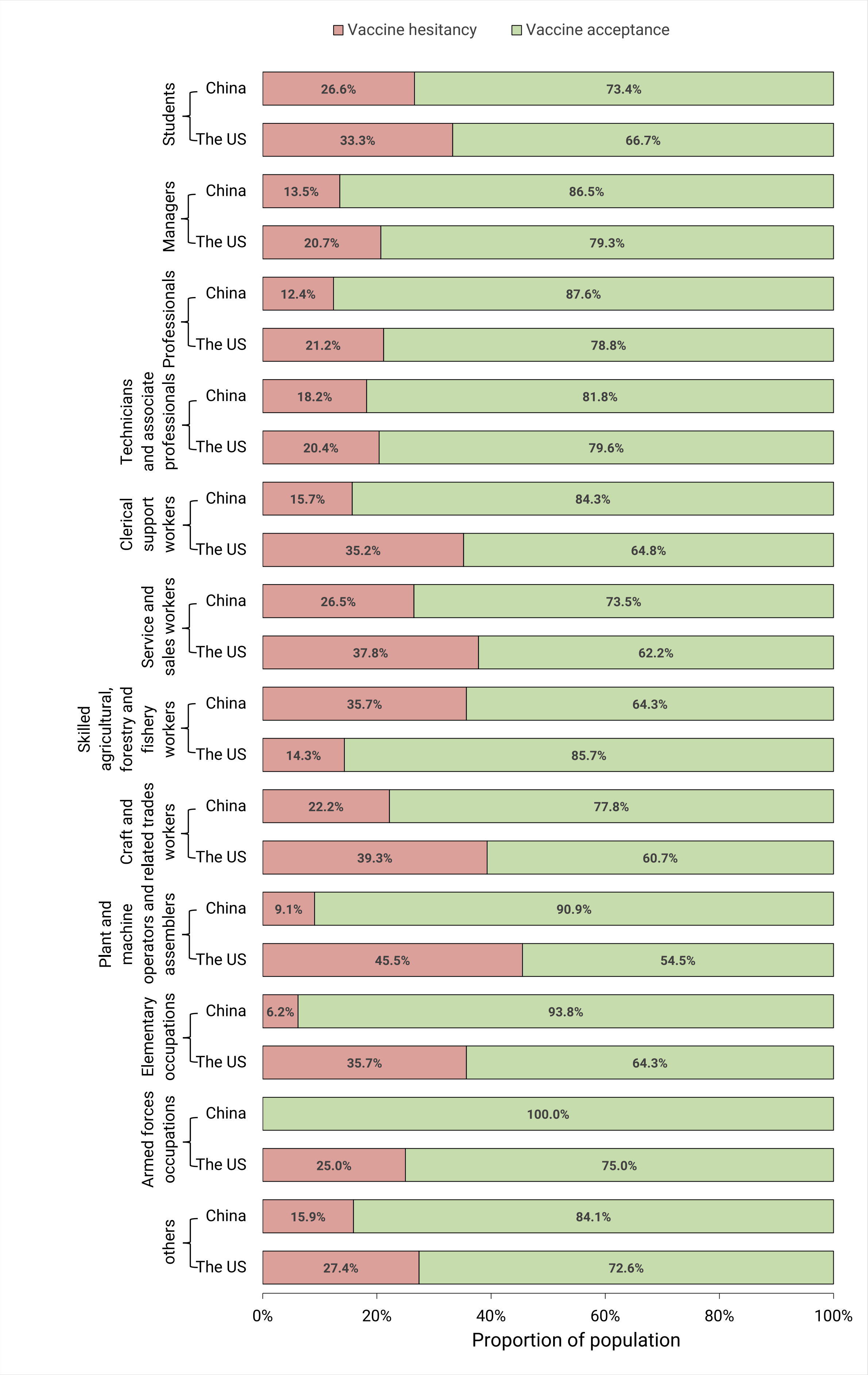

**Fig S4. COVID-19 vaccination acceptance in China before propensity score matching**

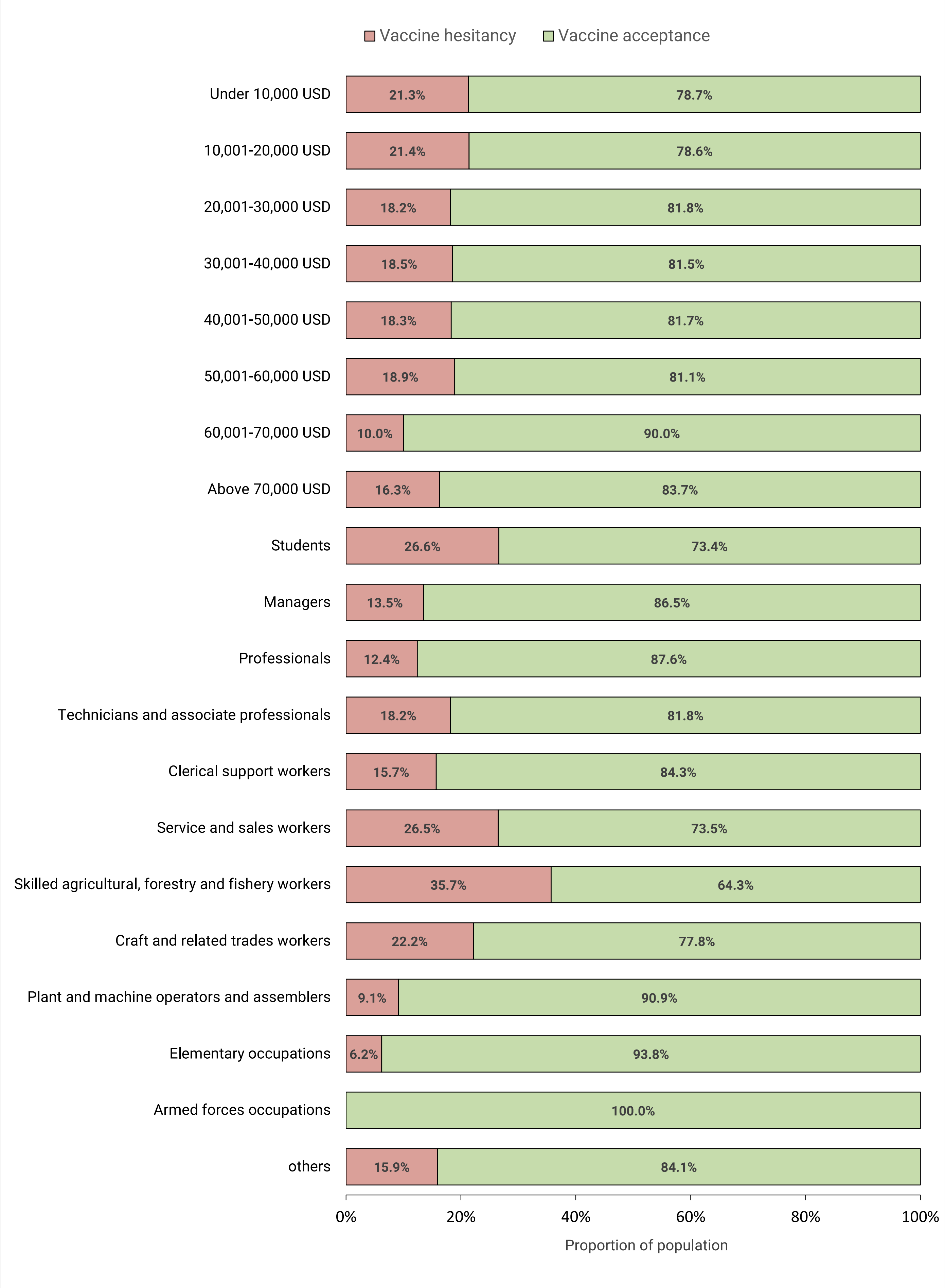

*(continued)*

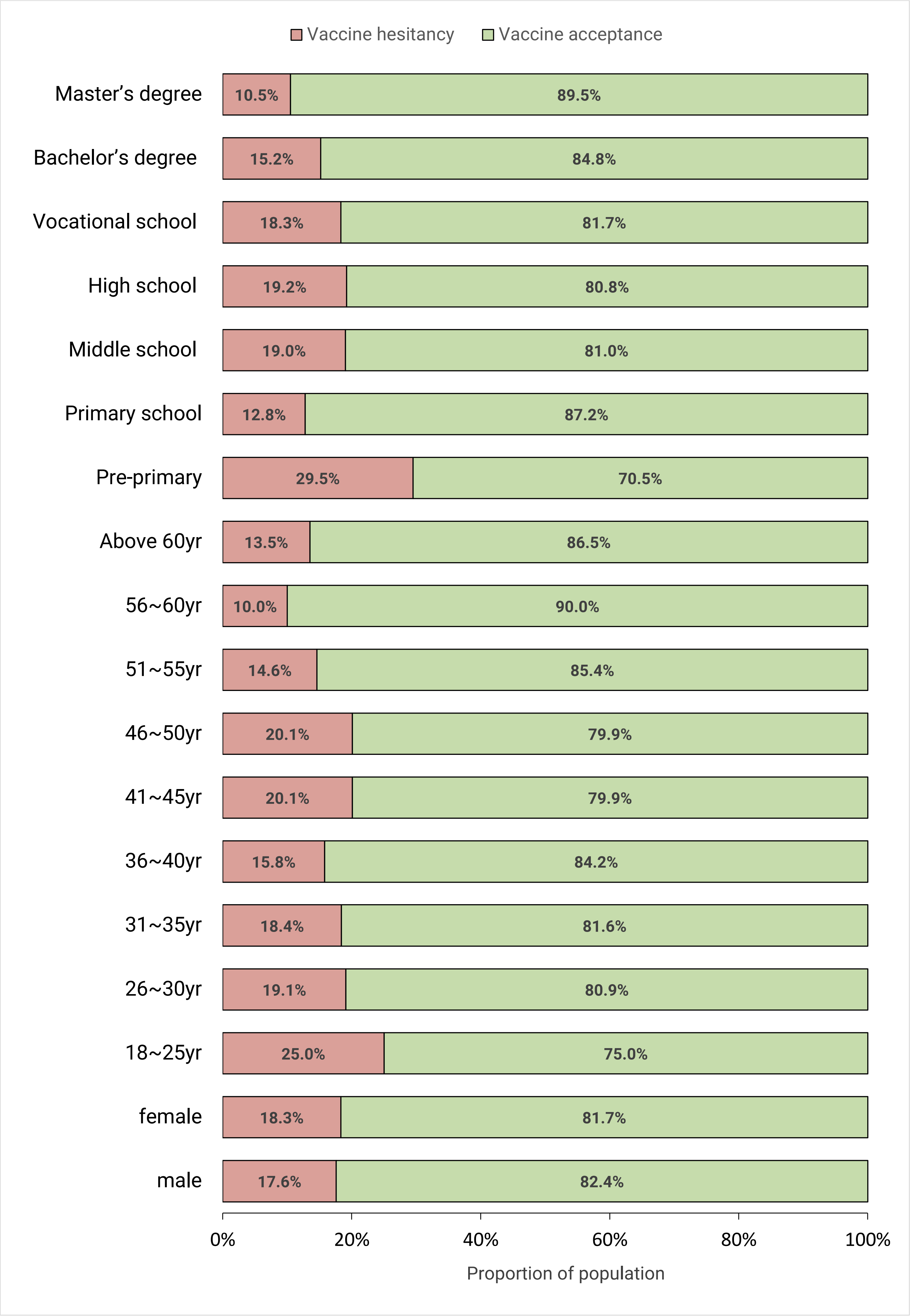

Vaccine hesitancy
Vaccine acceptance
Master’s degree
10.5%
89.5%
Bachelor’s degree
15.2%
84.8%
Vocational school
18.3%
81.7%
High school
19.2%
80.8%
Middle school
19.0%
81.0%
Primary school
12.8%
87.2%
Pre-primary
29.5%
70.5%
Above 60yr
13.5%
86.5%
56~60yr
10.0%
90.0%
51~55yr
14.6%
85.4%
46~50yr
20.1%
79.9%
41~45yr
20.1%
79.9%
36~40yr
15.8%
84.2%
31~35yr
18.4%
81.6%
26~30yr
19.1%
80.9%
18~25yr
25.0%
75.0%
female
18.3%
81.7%
male
17.6%
82.4%
0%
20%
40%
60%
80%
100%
Proportion of population

**Fig S5. COVID-19 vaccination acceptance in the US before propensity score matching**

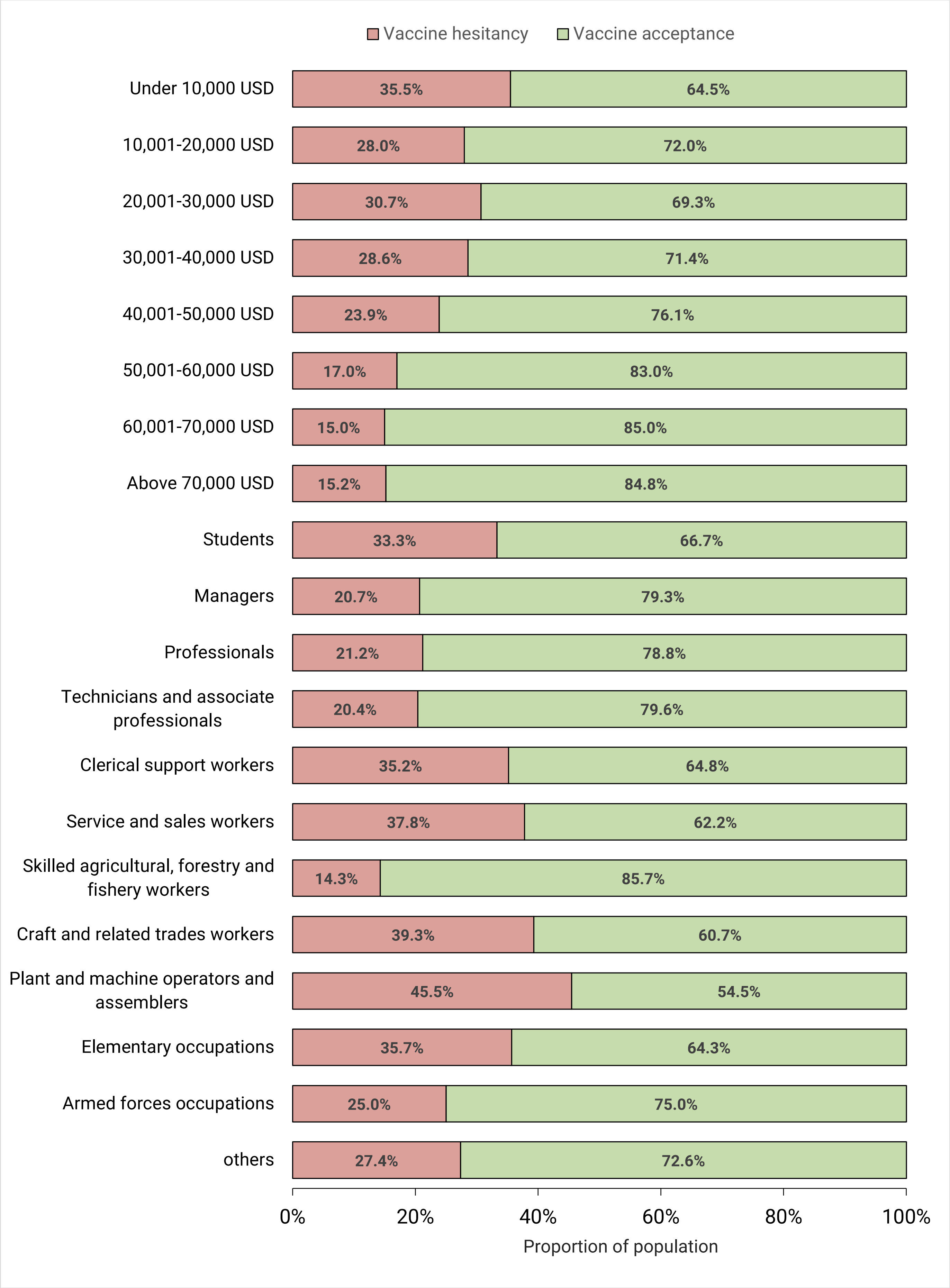

*(Continued)*

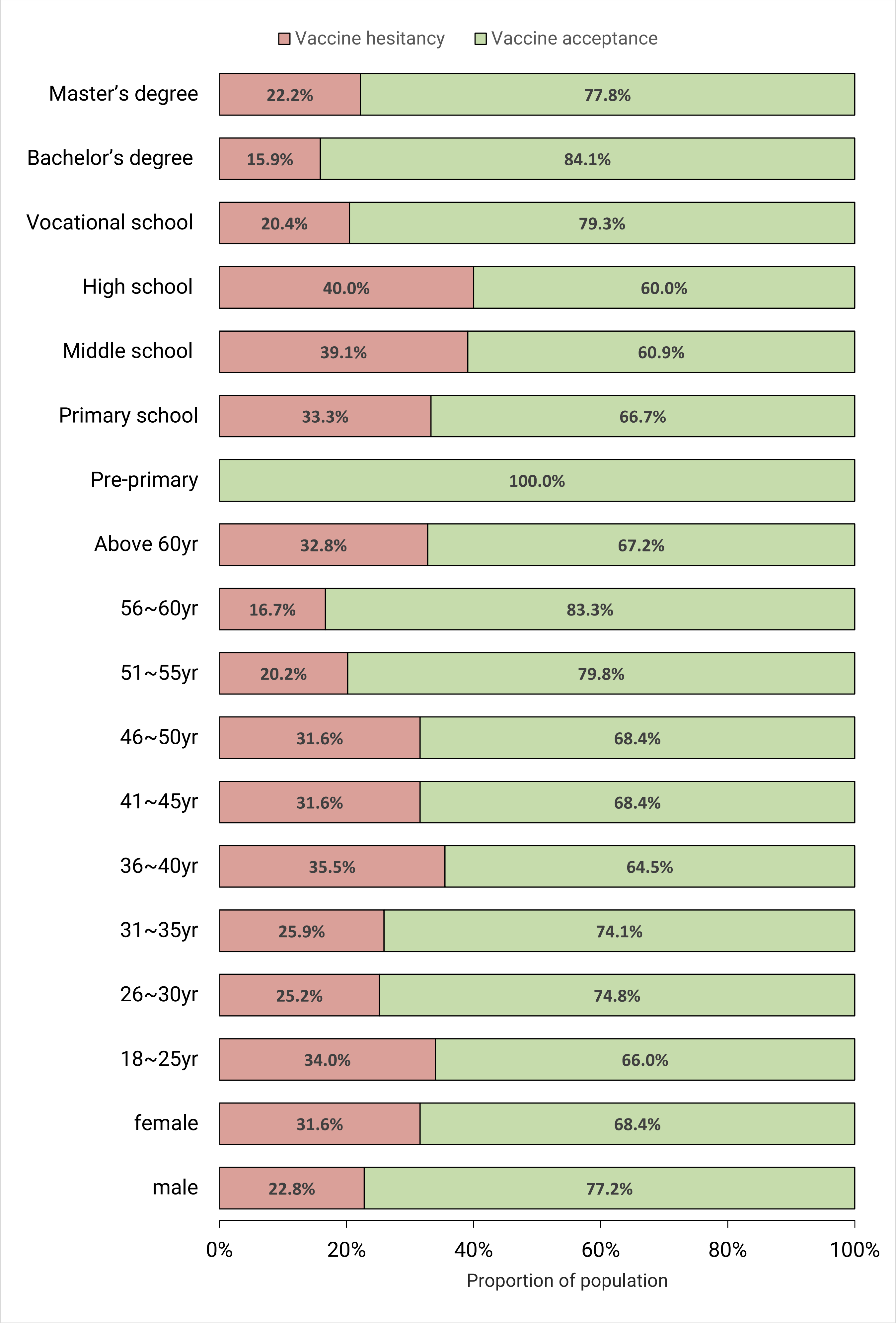
Vaccine hesitancy
Vaccine acceptance
Master's degree
22.2%
77.8%
Bachelor's degree
15.9%
84.1%
Vocational school
20.4%
79.3%
High school
40.0%
60.0%
Middle school
39.1%
60.9%
Primary school
33.3%
66.7%
Pre-primary
100.0%
Above 60yr
32.8%
67.2%
56~60yr
16.7%
83.3%
51~55yr
20.2%
79.8%
46~50yr
31.6%
68.4%
41~45yr
31.6%
68.4%
36~40yr
35.5%
64.5%
31~35yr
25.9%
74.1%
26~30yr
25.2%
74.8%
18~25yr
34.0%
66.0%
female
31.6%
68.4%
male
22.8%
77.2%
0%
20%
40%
60%
80%
100%
Proportion of population

**Fig S6. COVID-19 vaccination preference comparison between China and the US depending on sex after propensity score matching**

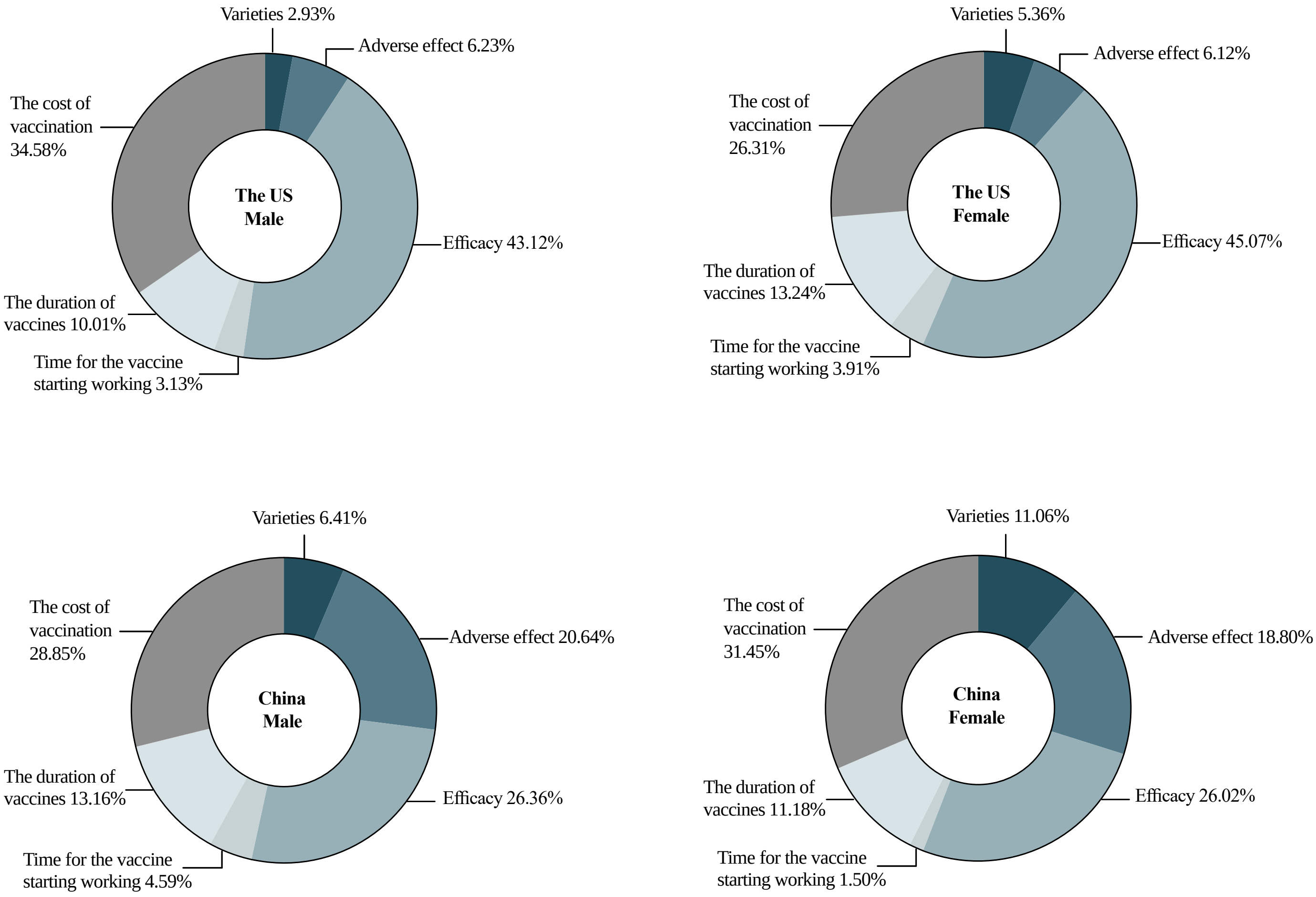

**Table S1. Behavioral and psychological results**

| | | Pre-PSM | | | Post-PSM | | |
|---|---|---|---|---|---|---|---|
| Items | | China | The US | *P* value | China | The US | *P* value |
| ***Past immunization behaviour/ adverse events*** | | (N=5374) | (N=3701) | | (N=1240) | (N=1240) | |
| “I have delayed getting a vaccine shot once (or more than once) for reasons other than illness or allergy." | Yes | 567 (10.6%) | 1246 (33.7%) | <0.001 | 152 (12.3%) | 475 (38.3%) | <0.001 |
| | No | 4807 (89.4%) | 2455 (66.3%) | | 1088 (87.7%) | 765 (61.7%) | |
| "I have once decided not to get a shot of vaccine for reasons other than illness or allergy?” | Yes | 528 (9.8%) | 1273 (34.4%) | <0.001 | 124 (10.0%) | 477 (38.5%) | <0.001 |
| | No | 4846 (90.2%) | 2428 (65.6%) | | 1116 (90.0%) | 763 (61.5%) | |
| “I have events in the past that discouraged me from getting a vaccine(s) for myself or my families?” | Yes | 363 (6.8%) | 1000 (27.0%) | <0.001 | 93 (7.5%) | 375 (30.2%) | <0.001 |
| | No | 5011 (93.2%) | 2701 (73.0%) | | 1147 (92.5%) | 865 (69.8%) | |
| Did you ever experience an AEFI (adverse event following immunization)? | Yes | 580 (10.8%) | 752 (20.4%) | <0.001 | 148 (12.0%) | 252 (20.3%) | <0.001 |
| | No | 4776 (89.2%) | 2937 (79.6%) | | 1089 (88.0%) | 987 (79.7%) | |
| ***Cues to action*** | | | | | | | |
| “I am recommended by a doctor to get COVID-19 vaccination.” | Yes | 1763 (32.8%) | 1968 (53.2%) | <0.001 | 395 (31.9%) | 622 (50.2%) | <0.001 |
| | No | 3611 (67.2%) | 1733 (46.8%) | | 845 (68.1%) | 618 (49.8%) | |
| “I am recommended by the local health board to get COVID-19 vaccination.” | Yes | 1995 (37.1%) | 2312 (62.5%) | <0.001 | 444 (35.8%) | 736 (59.4%) | <0.001 |
| | No | 3379 (62.9%) | 1389 (37.5%) | | 796 (64.2%) | 504 (40.6%) | |
| “I heard that my friends/families are being vaccinated.” | Yes | 2370 (44.1%) | 2572 (69.5%) | <0.001 | 502 (40.5%) | 803 (64.8%) | <0.001 |
| | No | 3004 (55.9%) | 1129 (30.5%) | | 738 (59.5%) | 437 (35.2%) | |
| **Likert Scale** | | | | | | | |
| **Perceived knowledge (source of information)** | | | | | | | |
| “I feel I get enough information about COVID-19 vaccines and their safety.” | | 5.1 (1.7) | 5.1 (1.8) | 0.36 | 5.2 (1.6) | 5.0 (1.9) | 0.002 |
| “The information I receive about COVID-19 vaccines from the vaccine program is reliable and trustworthy.” | | 5.3 (1.6) | 5.0 (1.9) | <0.001 | 5.3 (1.5) | 4.9 (1.9) | <0.001 |
| “I trust the information I receive about COVID-19 vaccines.” | | 5.2 (1.6) | 4.1 (2.0) | <0.001 | 5.3 (1.6) | 4.1 (2.0) | <0.001 |
| **Perceived severity** | | | | | | | |
| “I think there is a great chance for me to be infected with COVID-19.” | | 3.3 (2.2) | 4.5 (1.8) | <0.001 | 3.6 (2.2) | 4.5 (1.8) | <0.001 |
| “I think a COVID-19 infection would be a serious threat to health.” | | 5.8 (1.6) | 5.1 (1.9) | <0.001 | 5.8 (1.6) | 5.1 (1.8) | <0.001 |
| **Perceived benefits** | | | | | | | |
| “Getting COVID-19 vaccines is a good way to protect myself from COVID-19.” | | 5.7 (1.5) | 5.4 (1.9) | <0.001 | 5.7 (1.4) | 5.2 (2.0) | <0.001 |
| “Getting COVID-19 vaccines is a good way to prevent COVID-19 infection spread by patients.” | | 5.5 (1.6) | 5.4 (1.8) | <0.001 | 5.6 (1.5) | 5.3 (1.9) | <0.001 |
| “Getting COVID-19 vaccines is a good way to protect my friends or families from infected by COVID-19." | | 5.7 (1.4) | 5.4 (1.8) | <0.001 | 5.7 (1.4) | 5.3 (1.9) | <0.001 |
| **Perceived risks and barriers** | | | | | | | |
| “I am afraid that COVID-19 vaccine may, in fact, cause me to get infected with COVID-19.” | | 3.9 (2.1) | 3.0 (2.1) | <0.001 | 4.1 (2.1) | 3.2 (2.1) | <0.001 |

| | | | | | | |
|---|---|---|---|---|---|---|
| "I am concerned that I might have a serious side effect from a shot of COVID-19 vaccine." | 4.5 (1.9) | 4.0 (2.1) | <0.001 | 4.6 (1.8) | 4.2 (2.1) | <0.001 |
| "I am concerned that the production, storage, transportation, and unprofessional injection administration may cause the COVID-19 vaccines to be unsafe for taking." | 4.5 (1.9) | 3.8 (2.1) | <0.001 | 4.6 (1.9) | 3.9 (2.1) | <0.001 |
| **Perceived safety and efficacy of vaccines** | | | | | | |
| "I am concerned that taking the vaccines might not prevent COVID-19." | 4.5 (1.8) | 4.0 (2.0) | <0.001 | 4.5 (1.8) | 4.2 (2.0) | <0.001 |
| "It is better to develop immunity by getting vaccinated than by getting infected with COVID-19." | 5.5 (1.5) | 5.0 (2.0) | <0.001 | 5.6 (1.5) | 4.9 (2.1) | <0.001 |
| "I believe the COVID-19 vaccine is safe." | 5.3 (1.5) | 5.0 (1.9) | <0.001 | 5.3 (1.4) | 4.8 (2.0) | <0.001 |
| **General attitudes and trust for vaccines** | | | | | | |
| "In general, prevention is better than cure." | 5.9 (1.4) | 5.4 (1.7) | <0.001 | 5.9 (1.3) | 5.3 (1.7) | <0.001 |
| "In general, vaccination is effective in preventing diseases." | 5.7 (1.4) | 5.6 (1.6) | 0.033 | 5.7 (1.3) | 5.5 (1.6) | <0.001 |
| **Socio-cultural factors** | | | | | | |
| "I am able to openly discuss my concerns about COVID-19 vaccine shots with my doctor." | 5.4 (1.5) | 5.0 (1.9) | <0.001 | 5.4 (1.4) | 4.9 (1.9) | <0.001 |
| "I feel confident that the health center or doctor's office will have the COVID-19 vaccine when I need to take it." | 5.5 (1.5) | 5.5 (1.7) | 0.41 | 5.5 (1.5) | 5.4 (1.7) | 0.004 |
| "I trust that my government is making decisions in my best interest with respect to what COVID-19 vaccines are provided." | 5.4 (1.6) | 4.6 (1.9) | <0.001 | 5.4 (1.6) | 4.5 (1.9) | <0.001 |
| "I think COVID-19 vaccines are more important for boys/men" or "I think COVID-19 vaccines are more important for girls/women." | 5.7 (1.4) | 4.5 (1.9) | <0.001 | 5.7 (1.4) | 4.4 (2.0) | <0.001 |
| "I decide to take/not to take the COVID-19 vaccine because of religious or cultural reasons." | 4.0 (2.2) | 3.1 (2.1) | <0.001 | 4.2 (2.1) | 3.1 (2.0) | <0.001 |
| "I believe that people are risking their health or the health of the society if they do not take a COVID-19 vaccine." | 3.4 (2.3) | 2.4 (2.0) | <0.001 | 3.5 (2.3) | 2.5 (2.1) | <0.001 |
| "I think it is important for everyone to get the recommended COVID-19 vaccines for themselves." | 5.3 (1.6) | 4.9 (2.0) | <0.001 | 5.3 (1.6) | 4.7 (2.1) | <0.001 |
| "It's important for me to spend more than one hour in travel time to get a COVID-19 vaccine." | 5.7 (1.4) | 5.1 (2.0) | <0.001 | 5.7 (1.4) | 4.9 (2.0) | <0.001 |
| "I believe that COVID-19 vaccine producers care about my health more than their profit." | 5.7 (1.5) | 3.6 (2.1) | <0.001 | 5.7 (1.5) | 3.5 (2.2) | <0.001 |

**Table S2. The attributes and levels of vaccine acceptance and preference.**

| Attributes | Attributes' description | Levels |
|---|---|---|
| Vaccine varieties | Different varieties of vaccines developed in different countries | mRNA; adenovirus vector vaccines; Inactivated vaccine |
| Adverse effect | Extent of adverse effect after getting vaccinated | very mild; mild; moderate |
| Efficacy | The efficacy of vaccines that protect the vaccinators from getting infected with COVID-19 | 55%; 65%; 75%; 85%; 95% |
| Time for the vaccine starting working | Time period from getting vaccinated to the vaccine starts working | 5 days; 10 days; 15 days; 20 days |
| The duration of vaccine works | Time period from the vaccines starting to work to its invalidation | 5 months; 10 months; 15 months; 20 months |
| The cost of vaccination | The cost of whole vaccination process | $0; $50; $100; $150; $200 |

**Table S3. The Pearson Correlation matrix between the baseline characteristics and acceptance**

| Variables | -1 | -2 | -3 | -4 | -5 | -6 | -7 | -8 | -9 | -10 | -11 | -12 | -13 | -14 | -15 | -16 | -17 | -18 | -19 | -20 | -21 | -22 | -23 | -24 | -25 | -26 |
|---|---|---|---|---|---|---|---|---|---|---|---|---|---|---|---|---|---|---|---|---|---|---|---|---|---|---|
| (1) country | 1 | | | | | | | | | | | | | | | | | | | | | | | | | |
| (2) acceptance | 0.076* | 1 | | | | | | | | | | | | | | | | | | | | | | | | |
| (3) acceptance under social cues | 0.094* | 0.783* | 1 | | | | | | | | | | | | | | | | | | | | | | | |
| (4) sex | 0.031* | -0.061* | -0.047* | 1 | | | | | | | | | | | | | | | | | | | | | | |
| (5) age | 0.061* | 0.02 | 0.013 | -0.026 | 1 | | | | | | | | | | | | | | | | | | | | | |
| (6) education | -0.264* | 0.062* | 0.051* | -0.004 | -0.264* | 1 | | | | | | | | | | | | | | | | | | | | |
| (7) occupation | -0.099* | -0.049* | -0.045* | 0.039* | 0.321* | -0.286* | 1 | | | | | | | | | | | | | | | | | | | |
| (8) Annual earning | -0.560* | 0.018 | -0.003 | -0.098* | 0.017 | 0.420* | -0.119* | 1 | | | | | | | | | | | | | | | | | | |
| (9) ever_infected with COVID-19 | 0.264* | -0.025 | -0.011 | 0.050* | 0.051* | -0.109* | 0.014 | -0.144* | 1 | | | | | | | | | | | | | | | | | |
| (10) acquaintance ever infected with COVID-19 | 0.690* | 0.015 | 0.02 | -0.019 | 0.026 | -0.193* | -0.060* | -0.414* | 0.383* | 1 | | | | | | | | | | | | | | | | |
| (11) marital status | 0.028* | -0.043* | -0.051* | 0.014 | 0.378* | -0.120* | 0.189* | 0.062* | 0.064* | 0.066* | 1 | | | | | | | | | | | | | | | |
| (12) source_health~r | -0.007 | 0.066* | 0.068* | 0.014 | -0.051* | 0.096* | -0.053* | 0.039* | -0.036* | -0.026 | -0.041* | 1 | | | | | | | | | | | | | | |
| (13) source_PH_dep~t | -0.069* | 0.068* | 0.080* | 0.065* | -0.022 | 0.088* | -0.024 | 0.077* | 0.016 | -0.106* | -0.053* | 0.270* | 1 | | | | | | | | | | | | | |
| (14) source_news | 0.102* | 0.068* | 0.074* | 0.059* | 0.033* | 0.003 | 0.007 | -0.047* | 0.089* | 0.039* | -0.063* | -0.015 | 0.098* | 1 | | | | | | | | | | | | |

| | | | | | | | | | | | | | | | | | | | | | | | | | | |
|---|---|---|---|---|---|---|---|---|---|---|---|---|---|---|---|---|---|---|---|---|---|---|---|---|---|---|
| (15) source_social~a | 0.102* | 0.036* | 0.042* | 0.016 | -0.161* | 0.047* | -0.082* | -0.063* | -0.008 | 0.057* | -0.085* | 0.080* | 0.069* | 0.156* | 1 | | | | | | | | | | | |
| (16) source_friend~y | 0.062* | 0.022 | 0.034* | 0.044* | 0.013 | -0.043* | 0.026 | -0.049* | 0.022 | 0.036* | 0.018 | 0.107* | 0.069* | 0.126* | 0.327* | 1 | | | | | | | | | | |
| (17) source_Employer | -0.075* | 0.026 | 0.027* | -0.022 | -0.011 | 0.061* | 0.004 | 0.110* | -0.033* | -0.068* | 0.036* | 0.139* | 0.099* | 0.004 | 0.105* | 0.191* | 1 | | | | | | | | | |
| (18) source_ad | 0.097* | 0.033* | 0.023 | -0.018 | 0.022 | -0.033* | 0.001 | -0.059* | 0.036* | 0.069* | 0.026 | 0.210* | 0.160* | 0.092* | 0.179* | 0.226* | 0.291* | 1 | | | | | | | | |
| (19) source_other | -0.068* | -0.043* | -0.059* | -0.011 | -0.001 | 0.009 | 0.063* | 0.032* | -0.028* | -0.033* | -0.001 | -0.042* | -0.023 | -0.079* | -0.055* | -0.041* | -0.015 | -0.006 | 1 | | | | | | | |
| (20) delay_vaccine | 0.284* | 0.122* | 0.133* | 0.004 | 0.058* | -0.098* | -0.018 | -0.122* | 0.267* | 0.231* | -0.004 | -0.002 | 0.029* | 0.076* | -0.017 | -0.022 | -0.061* | 0 | -0.033* | 1 | | | | | | |
| (21) cancel_vaccine | 0.303* | 0.181* | 0.179* | -0.011 | 0.062* | -0.087* | -0.02 | -0.124* | 0.232* | 0.242* | -0.001 | -0.001 | 0.019 | 0.081* | -0.004 | -0.01 | -0.049* | -0.002 | -0.056* | 0.659* | 1 | | | | | |
| (22) discrouraging~s | 0.279* | 0.125* | 0.149* | 0.013 | 0.045* | -0.068* | 0.002 | -0.128* | 0.275* | 0.227* | -0.024 | -0.007 | 0.048* | 0.107* | -0.027 | -0.007 | -0.054* | 0.003 | -0.028* | 0.520* | 0.517* | 1 | | | | |
| (23) AEFI | 0.133* | 0.048* | 0.035* | 0.025 | 0.043* | -0.064* | 0.014 | -0.069* | 0.254* | 0.144* | -0.022 | -0.023 | -0.006 | 0.081* | -0.051* | -0.032* | -0.072* | -0.011 | -0.004 | 0.392* | 0.354* | 0.451* | 1 | | | |
| (24) doctor_recomm~d | 0.203* | -0.195* | -0.186* | 0.045* | -0.045* | -0.104* | -0.015 | -0.148* | 0.165* | 0.209* | -0.008 | -0.128* | -0.111* | 0.033* | 0.02 | -0.013 | -0.063* | -0.029* | 0.021 | 0.133* | 0.094* | 0.135* | 0.196* | 1 | | |
| (25) local_recommend | 0.249* | -0.171* | -0.167* | 0.030* | -0.039* | -0.109* | -0.026 | -0.159* | 0.149* | 0.244* | 0.006 | -0.119* | -0.142* | 0.025 | 0.027 | -0.007 | -0.082* | -0.023 | 0.008 | 0.139* | 0.116* | 0.126* | 0.178* | 0.744* | 1 | |
| (26) ff_vaccinated | 0.251* | -0.135* | -0.142* | -0.017 | -0.008 | -0.183* | 0.013 | -0.184* | 0.137* | 0.283* | 0.036* | -0.069* | -0.138* | -0.009 | 0.02 | -0.063* | -0.071* | 0.004 | 0.023 | 0.128* | 0.125* | 0.116* | 0.156* | 0.424* | 0.454* | 1 |
